# Ivabradine and Atrial Fibrillation Incidence: A Nested Matching Study

**DOI:** 10.1101/2025.01.10.25320367

**Authors:** Kibum Kim, Jasmeen Keur, Halie Anderson, Przemysław B. Radwański, Mark A. Munger

## Abstract

**Objective:** The objective of this real-world study was to quantify and project the cumulative incidence of atrial fibrillation (AF) associated with exposure to ivabradine (IVB) in naïve-AF heart failure (HF) patients.

**Methods:** A retrospective observational comparative study was performed using a health plan claims database. The new diagnosis of AF was compared between adult HF patients receiving IVB versus no IVB controls (CTRs) over 180-day follow-up period. Eligible IVB subjects were free from AF diagnosis before the IVB index date, which should be longer than 6 months. Incidence-density sampling was performed to select matched CTRs based on clinical characteristics, AF naïve status and time to be selected as a CTRs corresponding to the time to receive IVB from the initial HF diagnosis. The measure of IVB-AF association was tested using Cox Proportional Hazards Regression model.

**Results:** Of the 153 IVB and 4,494,305 CTRs meeting the AF-naïve HF status, the analytic cohort of 107 IVB and 321 matched controls were created. The groups were well matched for age: 52.9±11.3 (IVB) vs. 53.5±11.8, gender: 57% male, HFrEF diagnosis: 66.4%, co-morbidities: hypertension 81.3% and coronary artery disease event 38.3%, and goal-directed medical therapy: ß-adrenergic blockers 89.7%, ACEi/ARB/sacubitril/valsartan 79.4%, mineral corticoid antagonists 39.3%. The adjusted HR [95% CI] estimates was 7.293 [4.985-10.668]. The only variable that predicted AF incidence was the IVB prescription.

**Conclusions:** In HF AF-naive patients, analysis of healthcare claims point to a significant risk of AF receiving IVB.

## INTRODUCTION

Atrial fibrillation (AF) is the most common cardiac arrhythmia, with its prevalence steadily increasing worldwide. This rise can be attributed to the growing burden of cardiovascular disease, the increasing prevalence of metabolic disorders risk factors, and by drug-induced mechanisms.^1–2^ AF substantially contributes to morbidity, mortality, and medical expenditures.^3^ Health-care total costs including out-of-pocket, medical utilization, prescription drugs, and incremental costs are all substantially higher with AF versus non-AF patients.^4^ AF is preventable, early detection and appropriate treatment can substantially reduce AF-adverse outcomes.

Ivabradine (IVB) is a hyperpolarization-activated cyclic nucleotide-gated (HCN) channel blocker.^5^ The agent through inhibition of the cardiac pacemaker current (HCN-mediated funny current) [I_f_] current, causes spontaneous depolarization in the sino-atrial node that regulates heart rate.^6^ IVB is indicated for the treatment of NYHA FC II-IV chronic heart failure patients in sinus rhythm, in whom heart rate is ≥ 75 bpm in combination with goal-directed medical therapy (GDMT), including ß-adrenergic blockade or when ß-adrenergic blockade is contraindicated or not tolerated.^5^ IVB is associated with an increased incidence of AF through underlying gene mutations in HCN or pathophysiology AF triggers.^7^ In addition, heart failure measures and IVB pharmacology may modulate the effects of IVB to induce atrial fibrillation.^8–9^ In a recent meta-analysis of 13 randomized clinical trials with 37,533 patients, AF-incidence was significantly higher with IVB versus the control group, all with heart failure, [odds ratio (OR), 1.23; 95% confidence interval (CI), 1.08-1.41]. Subgroup analysis of left ventricular ejection fraction (LVEF) versus LVEF < 40% subgroups and small cumulative doses of IVB <100mg within versus large cumulative dose >300mg for 1-27.8 months were associated with higher AF IVB rates.^12^ In a separate meta-analysis with trial sequential analysis of 40,037 patients, it showed a 15% RR increase in atrial fibrillation with IVB treatment.^13^ However, Holter sub-studies from the BEAUTIFUL^14^ and SHIFT^15^ randomized clinical trials did not report a statistically significant increase in AF episodes, when comparing IVB to control patients at baseline, 1 month, or 8 months.

Based on these contrasting clinical trial findings, a large real-world study is warranted to determine atrial fibrillation incidence in patients taking IVB for heart failure. To this end, this real-world retrospective, nested-matched study was undertaken to assess the incidence of AF episodes with IVB compared to no IVB exposure.

## METHODS

### Data and Cohort Selection

A retrospective nested-matched study was performed using administrative claims from MarketScan^®^ Commercial Claims and Medicare Supplemental databases (Merative L.P., Ann Arbor, MI). The database consists of both medical and pharmacy claims for individuals covered by employment-based health plans. Each service encounter and procedure record are adjudicated by diagnosis codes. We utilized data from January 2009 through December 2021. Any information that can assist in identifying patients was removed by the data vendor prior to the investigators access to the data, making it impossible to link the data back to a specific individual. Using the databases were deemed exempt from the human subject research review by the University of Illinois Chicago Institutional Review Board.

The patient population was defined by individuals with diagnosis of heart failure (HF), which has been broadly considered as an indication of IVB and was defined by the presence of the International Classification of Disease 9^th^ or 10^th^ Revision Clinical Modification (ICD-9-CM 428.x or ICD-10-CM I50.x, and all subcodes) at any diagnosis position of the encounter. The date of the first HF diagnosis, following a 180-day HF-free baseline period during which patients were continuously enrolled without any coverage gaps, was designed as the “first HF date”. This date marked the start of patient follow-up to define exposure.

The first ever IVB dispensing was labelled as the Index exposure. To become eligible, patients had to be continuously enrolled in the database from the First HF. Eligible individuals were free from IVB exposure at any time prior to the Index exposure date, including the HF baseline period. If patients had a record of cardiac dysrhythmias defined by ICD-9-CM 427.x or ICD-10-CM I48.x before index exposure or during the HF baseline period, we excluded them from the research cohort. Only adult (≥ 18 years to no upper age limit) patients were selected as eligible IVB patients.

We used an incidence density sampling approach to select controls who were free from the exposure IVB while they were naïve to AF. Of the patients with HF, those who had not been exposed to IVB nor underwent the onset of AF until the corresponding IVB patient received IVB were selected as potential controls. The time from the First HF to the IVB date of the corresponding IVB patients were added to the control patients First HF date, which was flagged as “assigned Index date” for the controls. In theory, patients who received IVB later in the follow-up period could have a chance to be selected as a control prior to their index exposure.

For such cases, patient follow-up was supposed to end with exposure to IVB.

### Matching and Outcome assessment

The IVB – control matching was performed based on the patient characteristics collected over the 180-day period on or before either index exposure or assigned control date.

Demographics and patient characteristics assessed during the baseline period included grouped age (<45, 45-64, ≥ 65 years), sex, year of the index exposure date or corresponding assigned index date, acute coronary syndrome or acute myocardial infarction, hypertension, hyperlipidemia, diabetes, record of hospital admission with HF diagnosis at any diagnosis position of the admission summary, and presence of heart failure with reduced ejection fraction (HFrEF). To reduce the concern on the unobserved or missing diagnosis codes that potentially became a confounders in the IVB-AF association, we also matched the baseline medication classes that indicates the severity or stages of HF progression, including angiotensin-converting enzyme inhibitors, angiotensin receptor blockers, and sacubitril/valsartan under the category of renin-angiotensin inhibitors (RAS), cardiac beta-blockers, mineralocorticoid receptor antagonists, and sodium-glucose cotransporter-2 inhibitors. We performed a one-by-three matching with replacement, and each of the listed patient-level characteristics were exactly matched between the IVB and controls (Figure 1).

**Figure 1.**
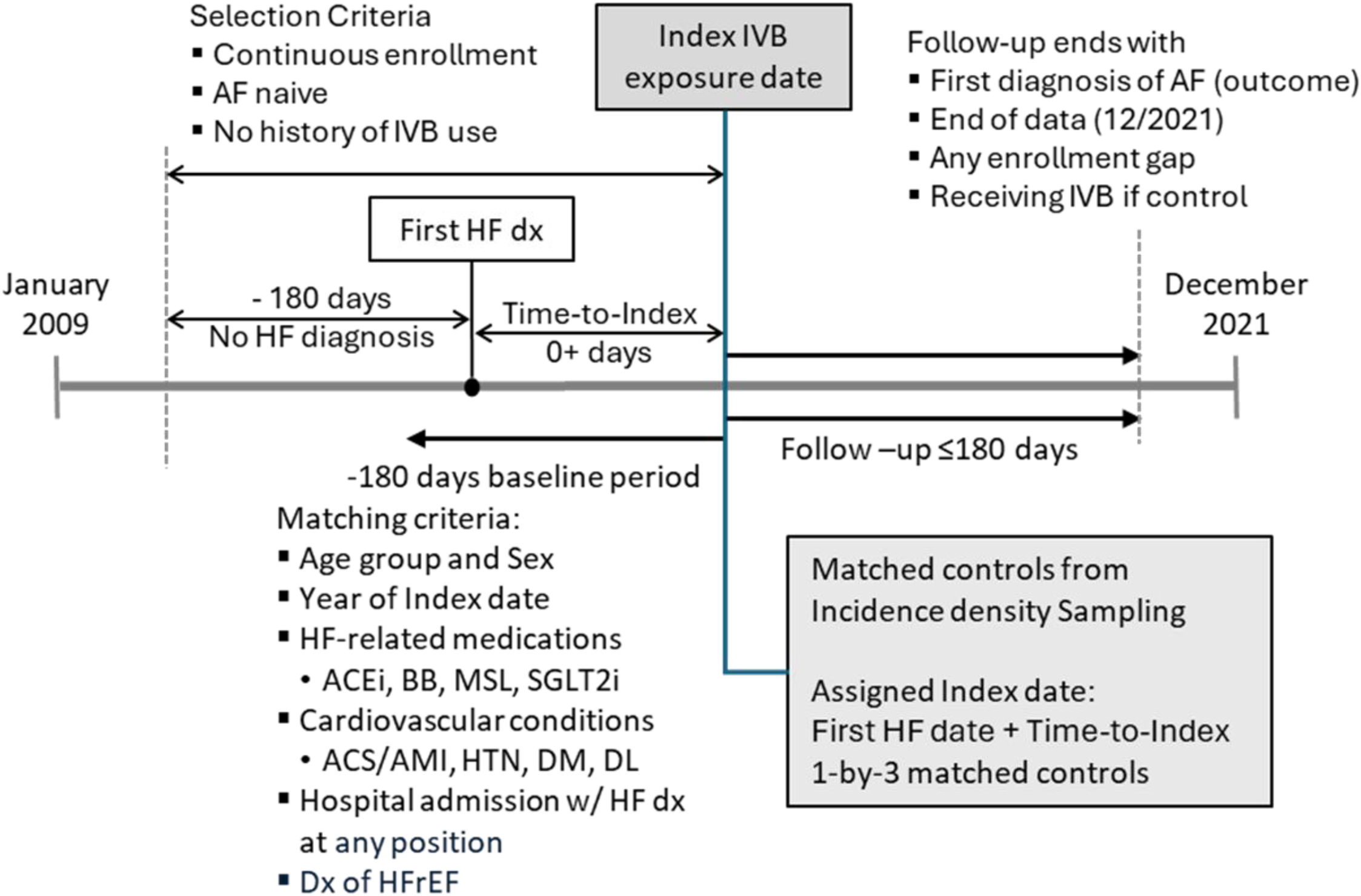
**Cohort Selection and Matching**

We assessed the onset of AF as the outcome of this study over the 180-day follow-up period from the index exposure or assigned index date. The outcome of this study was the onset of AF defined by the diagnosis code (ICD-9-CM 427.31, ICD-10-CM I48.0 or I48.1) at any position that adjudicate the medical care encounter. For the time-to-event analysis, the patient follow-up ended with the onset of AF, end of continuous enrollment with any gap, or end of the 180-day follow-up period.

### Statistical Analysis

Patients characteristics, after incidence density sampling but before matching and after the completion of 1-by-3 matching, were compared between the IVB and controls using descriptive statistics followed by bivariate analysis. The summary statistics for age in years were mean and standard deviation, which were compared using Student *t*-test. All other categorical variables were summarized using frequency and percentage. We used Chi-square test of independence to test the statistical difference in the patient characteristics between the IVB and controls.

Time to AF onset from the index date was summarized and presented using cumulative incidence from Kaplan-Meier product limit estimates. While matched patients were followed until the onset of AF up to 180 days from the index date, following censoring criteria were applied: end of 180-day follow up period, end of study data (December 2021), any enrollment gap, and receiving IVB if a patient was enrolled in the cohort as a control. The comparative outcome assessment was performed using Cox Proportional Hazards Regression model from which hazard ratio (HR) and 95% confidence intervals (95% CI) of the onset of AF for IVB vs. control cohorts.

## RESULTS

Of all the patients with the diagnosis of HF from the database, 153 IVB patients and 4,494,305 controls from incidence density sampling met the continuous enrollment and AF-naïve criteria before either IVB or assigned index date. Before matching patient characteristics, IVB patients were younger than the controls with respective mean (SD) year ages of 52.5 (12.2) and 61.2 (16.7). While middle-aged (45 – 64 years) accounted for 66.7% of the IVB patients, the corresponding proportion among the controls was 49.1%. IVB patients had elevated risks of cardiovascular complications compared to the controls with the respective proportion of ACS/AMI history, hyperlipidemia, HFrEF, and admission with HF diagnosis of 19.6% (vs. 8.9%), 60.8% (vs. 56.9%), 67.3% (vs. 24%), 51.0% (vs. 29.2%). The elevated risk of cardiovascular complications was also shown in the more use of cardiovascular medications with the respective proportions of RASi, BB, MRA and SGLT2i use of 76.5% (vs. 49.7%), 91.5% (vs. 44.6%), 46.4% (vs. 8.4%) and 8.5% (vs. 2.5%). Summary statistics and statistical significance test results are presented in the Table 1.

**Table 1.**
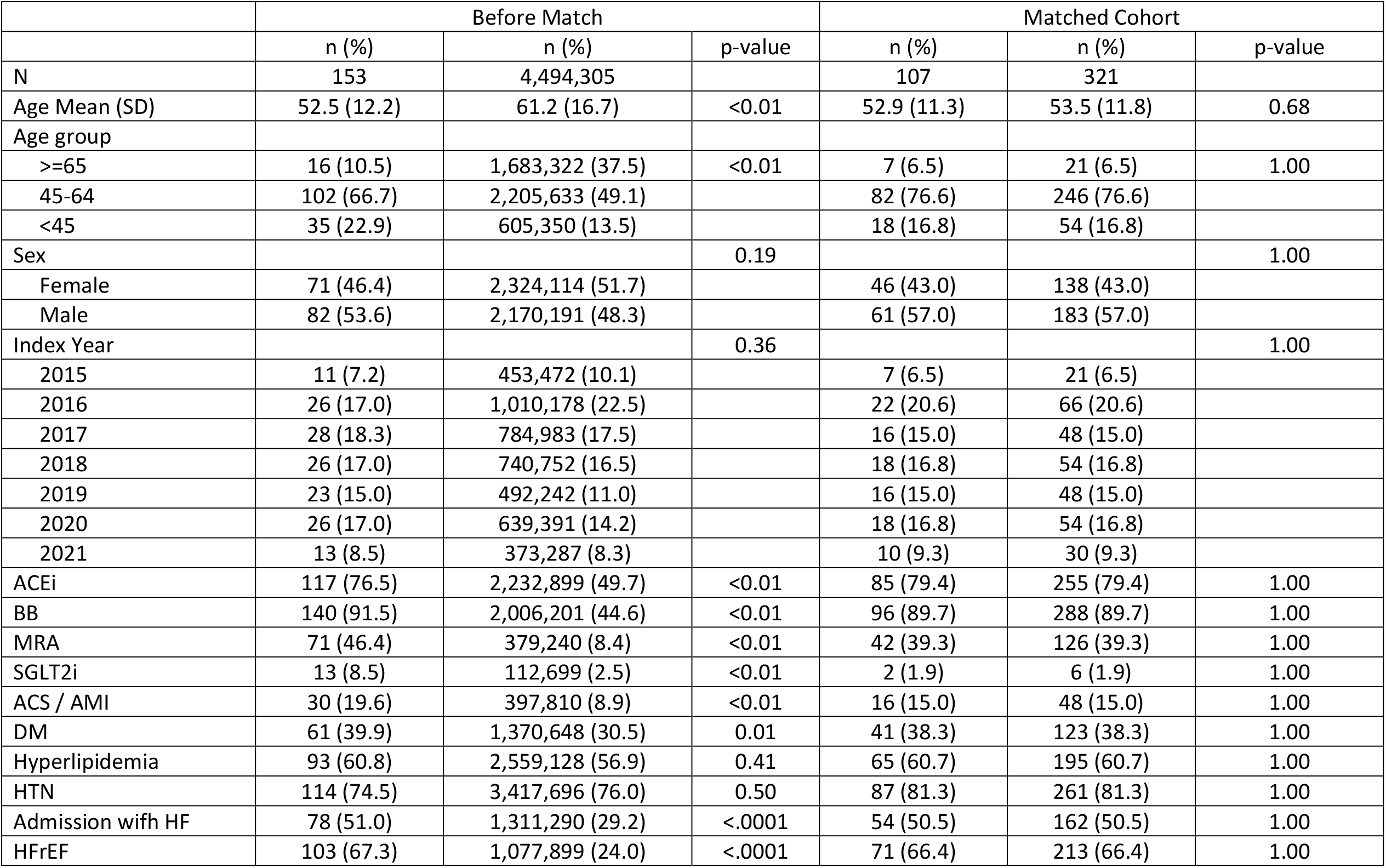
Baseline Characteristics.

|  | Before Match |  |  | Matched Cohort |  |  |
| --- | --- | --- | --- | --- | --- | --- |
|  | n (%) | n (%) | p-value | n (%) | n (%) | p-value |
| N | 153 | 4,494,305 |  | 107 | 321 |  |
| Age Mean (SD) | 52.5 (12.2) | 61.2 (16.7) | <0.01 | 52.9 (11.3) | 53.5 (11.8) | 0.68 |
| Age group |  |  |  |  |  |  |
| >=65 | 16 (10.5) | 1,683,322 (37.5) | <0.01 | 7 (6.5) | 21 (6.5) | 1.00 |
| 45-64 | 102 (66.7) | 2,205,633 (49.1) |  | 82 (76.6) | 246 (76.6) |  |
| <45 | 35 (22.9) | 605,350 (13.5) |  | 18 (16.8) | 54 (16.8) |  |
| Sex |  |  | 0.19 |  |  | 1.00 |
| Female | 71 (46.4) | 2,324,114 (51.7) |  | 46 (43.0) | 138 (43.0) |  |
| Male | 82 (53.6) | 2,170,191 (48.3) |  | 61 (57.0) | 183 (57.0) |  |
| Index Year |  |  | 0.36 |  |  | 1.00 |
| 2015 | 11 (7.2) | 453,472 (10.1) |  | 7 (6.5) | 21 (6.5) |  |
| 2016 | 26 (17.0) | 1,010,178 (22.5) |  | 22 (20.6) | 66 (20.6) |  |
| 2017 | 28 (18.3) | 784,983 (17.5) |  | 16 (15.0) | 48 (15.0) |  |
| 2018 | 26 (17.0) | 740,752 (16.5) |  | 18 (16.8) | 54 (16.8) |  |
| 2019 | 23 (15.0) | 492,242 (11.0) |  | 16 (15.0) | 48 (15.0) |  |
| 2020 | 26 (17.0) | 639,391 (14.2) |  | 18 (16.8) | 54 (16.8) |  |
| 2021 | 13 (8.5) | 373,287 (8.3) |  | 10 (9.3) | 30 (9.3) |  |
| ACEi | 117 (76.5) | 2,232,899 (49.7) | <0.01 | 85 (79.4) | 255 (79.4) | 1.00 |
| BB | 140 (91.5) | 2,006,201 (44.6) | <0.01 | 96 (89.7) | 288 (89.7) | 1.00 |
| MRA | 71 (46.4) | 379,240 (8.4) | <0.01 | 42 (39.3) | 126 (39.3) | 1.00 |
| SGLT2i | 13 (8.5) | 112,699 (2.5) | <0.01 | 2 (1.9) | 6 (1.9) | 1.00 |
| ACS / AMI | 30 (19.6) | 397,810 (8.9) | <0.01 | 16 (15.0) | 48 (15.0) | 1.00 |
| DM | 61 (39.9) | 1,370,648 (30.5) | 0.01 | 41 (38.3) | 123 (38.3) | 1.00 |
| Hyperlipidemia | 93 (60.8) | 2,559,128 (56.9) | 0.41 | 65 (60.7) | 195 (60.7) | 1.00 |
| HTN | 114 (74.5) | 3,417,696 (76.0) | 0.50 | 87 (81.3) | 261 (81.3) | 1.00 |
| Admission with HF | 78 (51.0) | 1,311,290 (29.2) | <.0001 | 54 (50.5) | 162 (50.5) | 1.00 |
| HFrEF | 103 (67.3) | 1,077,899 (24.0) | <.0001 | 71 (66.4) | 213 (66.4) | 1.00 |

By matching patient demographics and risk indicators, we were able to create an analytic cohort consisting of 107 IVB patients and 321 matched controls. The matching process eliminates the patients with extremely high-risk from the IVB groups while selecting the patients with elevated risk profiles from the control groups. The mean (SD) year age for IVB and matched controls after matching was 52.9 (11.3) and 53.5 (11.8), respectively, with the proportion of middle-aged patients of 76.6% for both IVB and control groups. The proportion of HF-related medications and cardiovascular risk indicators were exactly matched (Table 1).

The Kaplan-Meier estimates from the matched cohort indicated that the cumulative incidence of AF at 180-days after the index date for IVB and controls were 55.4% and 10.1%, respectively. The hazard ratio estimate [95% confidence interval] from the cox-proportional hazard regression model was 7.293 [4.985 −10.668] (Figure 2).

**Figure 2.**
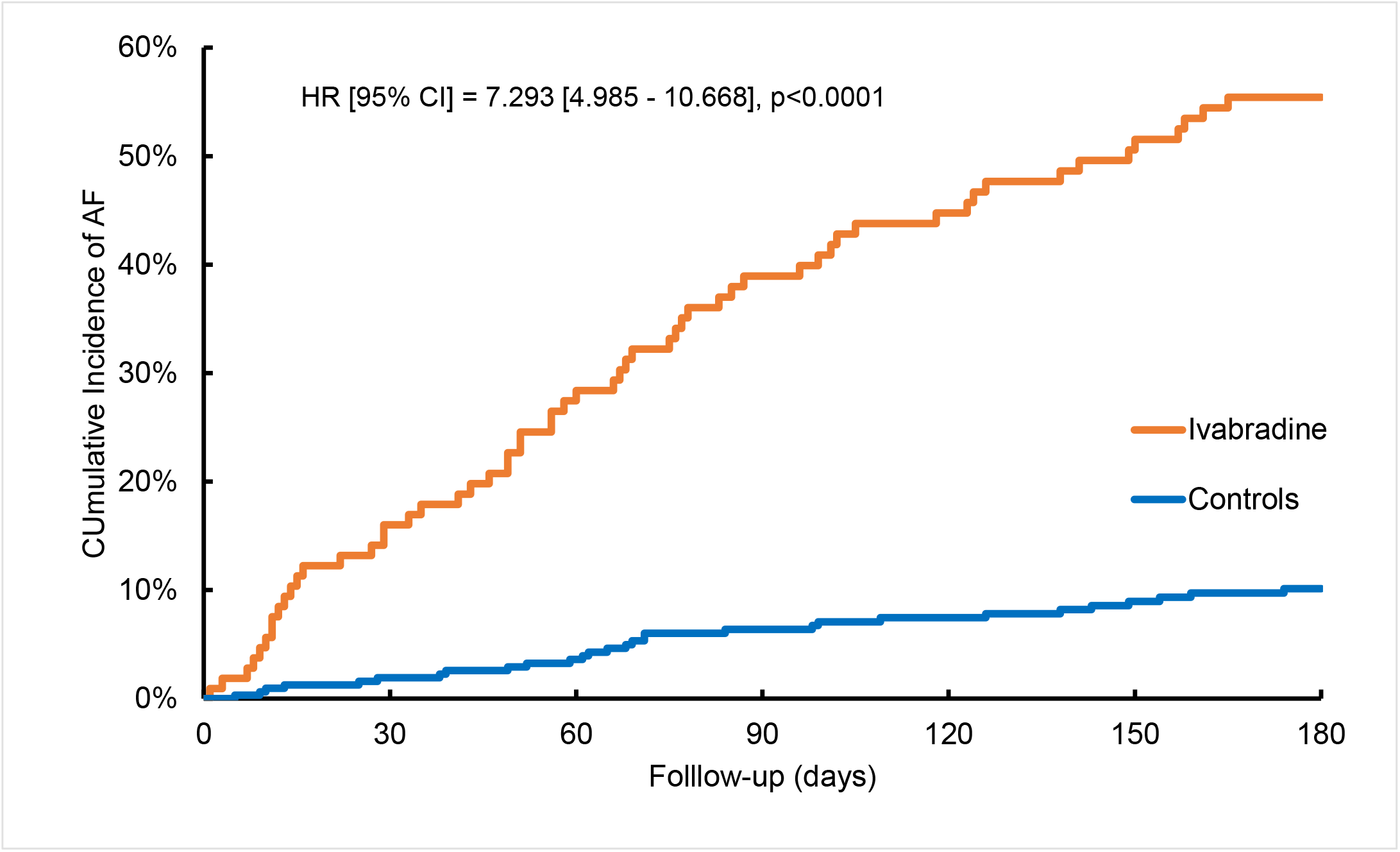
**Cumulative incidence of AF and Hazard Ratio estimate**

## DISCUSSION

In a nested-matching study from a United States-based commercial insurance and Medicare supplement claims database, AF-naïve patients exposed to IVB was associated with a substantial statistical increase in the cumulative incidence of AF when compared to no IVB exposure in heart failure patients. This association remained a significant event after adjusting for all confounding factors. On the backdrop of extensive use of beta-blockers and angiotensin converting enzyme in our study population, these results suggest AF incidence in a real-world data may be a more clinically important arrhythmogenic outcome than previously reported from clinical trials.^12–15^

IVB-associated AF-incidence may be an association more than a cause-effect relationship. However, regulation of heart rate by IVB occurs through blockade of the cardiac pacemaker current (HCN funny current) [I_f_].^16^ Pulmonary vein myocardial sleeves which are notable AF triggers have I_f_ currents.^17^ The IVB arrhythmogenic effect is seen immediately (Figure 2) suggesting that the effect is not dose-related, but potentially a pharmacologic association which is mediated by genetically or physiologically.^18^

In the previous important clinical trials of IVB for FDA approval^14–15^, AF was an exclusion criterion. In a multicenter, observational matched cohort study 887 patients prescribed IVB compared to 1115 with no IVB in symptomatic HFrEF with history of paroxysmal AF, IVB treatment was associated with an increased risk of the primary outcome of HF hospitalization and HF death (HR: 1.56; 95% CI: [1.40-1.75] and 1.67 [1.14-re2.44]).^19^ These combined with our current findings *may* provide further evidence for discussions, the need for further clinical trials to confirm the incidence in a general population, AF outcomes including self-termination, recurrence rate, termination post-discontinuation of IVB, the need for rhythm-control agents or ablation for control of symptoms, and potential reassessment of the role of IVB in HF treatment.

### Limitations

There are limitations to this study. It is a retrospective nested-matched analysis of healthcare claims. While the analytic approach did not rule out the possibility of the confounding by indication, the cumulative incidence of AF from the study gives a warning sign for AF risk. The accuracy of identifying patients and outcomes is subject to the accuracy of the diagnosis codes.

Using diagnosis codes for AF attained the positive predictive value ranged from 55% to 90%, raising questions on the accuracy of the estimated incidence. Nevertheless, the anticipated impact of the confounding would not be large enough to pay off the size and direction of the association. calculating hazard ratio, the impact of the coding accuracy would likely be similar between IVB and CTR groups, making any bias in the direction or strength of the association minimal. Lastly, general limitations of retrospective observational research including unobserved confounders, left censoring of patients before receiving IVB should also be considered.

## CONCLUSION

In HF AF-naive patients, analysis of healthcare claims supports a significant risk of developing AF when treated with IVB.

## Data Availability

All data will be available through the corresponding author upon request.

